# White matter abnormalities characterise the acute stage of sports-related mild Traumatic Brain Injury

**DOI:** 10.1101/2022.02.07.22270651

**Authors:** Remika Mito, Donna M. Parker, David F. Abbott, Michael Makdissi, Mangor Pedersen, Graeme D. Jackson

## Abstract

Sports-related concussion, a form of mild traumatic brain injury (mTBI), is characterised by transient disturbances of brain function. There is increasing evidence that subtle abnormalities drive functional brain changes in white matter microstructure, and diffusion MRI has been instrumental in demonstrating these white matter abnormalities *in vivo*. However, the reported location and direction of the observed white matter changes in mTBI are variable, likely attributable to the inherent limitations of the white matter models used. This cross-sectional study applies an advanced and robust technique known as fixel-based analysis to investigate fibre tract-specific abnormalities in professional Australian Football League players with a recent mTBI. We used the fixel-based analysis framework to identify common abnormalities found in specific fibre tracts in participants with an acute mTBI (≤ 12 days after injury; *n* = 14). We then assessed whether similar changes exist in subacute mTBI (> 12 days and < 3 months after injury; *n* = 15). The control group was 29 neurologically healthy control participants. We assessed microstructural differences in fibre density and fibre bundle morphology and performed whole-brain fixel-based analysis to compare groups. Subsequent tract-of-interest analyses were performed within five selected white matter tracts to investigate the relationship between the observed tract-specific abnormalities and days since injury and the relationship between these tract-specific changes with cognitive abnormalities. Our whole-brain analyses revealed significant increases in fibre density and bundle cross-section in acute mTBI when compared to controls. Acute mTBI showed even more extensive differences when compared to subacute mTBI than to controls. The fibre structures affected in acute mTBI included the corpus callosum, left prefrontal and left parahippocampal white matter. The fibre density and cross-sectional increases were independent of time since injury in acute mTBI, and were not associated with cognitive deficits. Overall, this study demonstrates that acute mTBI is characterised by specific white matter abnormalities, which are compatible with tract-specific cytotoxic oedema. These potential oedematous changes were absent in our subacute mTBI participants, suggesting that they may normalise within 12 days after injury, although subtle abnormalities may persist in the subacute stage. Future longitudinal studies are needed to elucidate individualised recovery after brain injury.

## Introduction

Over the past two decades, there has been rapidly growing interest in the short- and long-term pathophysiological effects of sports-related concussion, in large part, prompted by affected high-profile athletes and forced retirements across numerous different sporting codes. Sports-related concussion, or mild traumatic brain injury (mTBI), is characterised by a complex cascade of neurometabolic events, which can impact brain function for days to weeks and months^1^. Given the absence of macroscopic abnormalities on conventional structural MRI, mTBI is considered a functional disturbance, rather than structural injury^2,3^. In moderate to severe TBI, however, the functional disturbances that arise are thought to be a consequence of structural damage to axonal fibre pathways within large-scale brain networks^4,5^. Traumatic axonal injury is a hallmark pathological feature of TBI^6–9^, and despite the paucity of post-mortem studies in mild TBI^10^, there is now increasing consensus that subtle damage to white matter microstructure also underpins the functional deficits experienced in mTBI. Investigating subtle changes to white matter fibre structures *in vivo* could thus be valuable in the diagnosis and management of mTBI.

To this end, advances in human neuroimaging have resulted in an improved capability to examine tissue microstructure *in vivo*, and in particular, diffusion-weighted imaging (DWI) has provided a means to non-invasively examine white matter architecture. A particular form of DWI, known as diffusion tensor imaging (DTI), has been widely adopted by researchers to assess subtle white matter abnormalities in mTBI participants. DTI studies have identified abnormalities at various time-points following sports-related mTBI^11–16^. These studies have typically assessed changes to metrics such as fractional anisotropy and mean or radial diffusivity, which in more severe cases of TBI have been used as an *in vivo* proxy for traumatic axonal injury^17^. However, in mTBI, the subtle nature of neurophysiological effects means that white matter tract changes are challenging to assess. There has correspondingly been substantial variability in the direction of detected DTI-based changes, as well as the spatial location of these abnormalities across studies^18–22^.

A potential explanation for the inconsistencies across DTI studies may stem from limitations to the DTI model itself. While DTI is sensitive to subtle abnormalities to white matter microstructure, it is unable to model complex or crossing-fibre populations, which account for up to 90% of white matter imaging voxels^23^. In the presence of such crossing-fibre structures, the same underlying pathologies could drive both increases and decreases in DTI-based metrics^24–27^. For example, a loss of axons could result in decreased fractional anisotropy; however, in the presence of unaffected crossing fibre structures, loss of axons could result in increased anisotropy in that region^25,27^. Indeed, in the context of mTBI, both increases^11,13,28,29^ and decreases^30–32^ in fractional anisotropy have been reported. These conflicting findings, though they have been suggested to reflect the same pathophysiological changes^11^, can be difficult to interpret biologically, and as such, researchers have stopped short of recommending DTI as a diagnostic biomarker in the clinical setting.

Higher-order DWI models overcome some of the limitations inherent to the DTI model and may be critical for detecting subtle white matter abnormalities in mTBI without being confounded by the presence of crossing-fibre structures. While several models can estimate multiple fibre orientations within voxels^33–35^, one recent DWI analysis technique known as fixel-based analysis (FBA)^36^ also enables quantification of changes to specific fibre populations within voxels. FBA can be used to estimate: (i) differences in the density of fibres within a fibre bundle, (ii) differences in fibre bundle cross-section, and (iii) differences arising from a combination of density and cross-sectional changes^36,37^. By performing statistical analyses at each white matter ‘fixel’ (specific *<u>fi</u>*bre population within a vo*<u>xel</u>*), FBA can ascribe detected changes to specific white matter tracts and does not suffer from interpretability issues that can arise in crossing-fibre populations when using DTI^27,38^. Moreover, even in the absence of potential confounding factors that arise due to crossing-fibre populations, FBA metrics can offer greater biological interpretability, which could be highly valuable in the clinical setting in mTBI.

In this cross-sectional study, we applied FBA to investigate fibre tract-specific abnormalities in 29 professional Australian Football League players with a recent mTBI. These professional players were available to participate in the study only when they were unable to return to training and play. This was either after acute injury before return-to-play, or sub-acutely when they had persistent symptoms that prevented them from return-to-play. This study aims to investigate whether sports-related mTBI is characterised by abnormalities within specific white matter fibre tracts at these acute and subacute stages. We hypothesised that individuals with acute mTBI would exhibit tract-specific fibre density abnormality, and that these abnormalities would be less pronounced in participants at the subacute phase of injury. Additionally, we investigated the association of these fibre-specific abnormalities with time since injury and cognition in mTBI, hypothesising a decline of abnormality with greater time since injury and more apparent abnormality in those with greater cognitive symptomatology.

## Materials and methods

### Participants

Male participants (*n* = 38) were recruited from professional Australian Football League (AFL) teams in Australia between 2014 and 2019. AFL is a sport played at speed and with a high incidence of concussion, of up to 17.6 concussions per 1000 player hours^39–41^. All participants were recruited after having suffered a suspected mTBI (concussion). Concussion was diagnosed by experienced team doctors who were present at the time of injury. AFL team doctors have a consistent approach to the diagnosis and management of concussions due to ongoing research and education programs over the past 20 years^39,42^.

All concussed participants were assessed at the time of injury with the Sports Concussion Assessment Tool (SCAT3), and a sideline video review was used where available^43^. All concussed players had clinical signs and symptoms of concussion lasting 72 hours or longer. Participants were excluded from the study where a diagnosis of concussion was not clinically confirmed (*n* = 2). Anatomical MRI scans were reviewed by a neurologist (GJ). Participants were also excluded if they exhibited a focal injury or more severe TBI resulting in intracranial bleeding (e.g., subdural hematoma; *n* = 1). Participants were also excluded from the study if their MRI scan was more than 3 months after their concussion (*n* = 3), if they had been scanned using a different MRI scanner type or head coil (*n* = 1), or if their MRI acquisition was incomplete (*n* = 2).

In total, 29 male professional AFL footballers were included in the study. These 29 mTBI participants were categorised into two groups, based on the time delay from the concussion until MRI scan. Those participants who had their MRI scan ≤ 12 days since concussion were categorised into an acute mTBI group (*n* = 14), while those whose MRI scan was > 12 days since concussion were categorised into a subacute mTBI group (*n* = 15). Although acute concussive injury is commonly defined as injury within a 48-hour period^2^, researchers use various time periods to describe acute mTBI across studies^22^. Here, we use a cut-off period of 12 days in line with current AFL protocol that mandates a 12-day rest period following mTBI^44^.

Given that all mTBI participants were professional athletes with substantial training commitments, involvement in the study was possible when the footballers had not yet returned to training and competition. This meant that participants categorised into the subacute mTBI group tended to have persistent symptoms lasting beyond 12 days, preventing them from returning to play.

Healthy control participants were recruited for the study, or selected from previously acquired DWI control data obtained at our institute (*n* = 29). For each mTBI participant, a male control participant was selected who matched the mTBI participant in age.

All participants provided written informed consent before participating in the study. The study was approved by the human research ethics committees at the University of Melbourne (ID: 0830367) and at Austin Health (ID: 49573/2019).

### MRI data acquisition

All participants underwent an MRI scan at the Florey Institute of Neuroscience and Mental Health, which included high angular resolution diffusion-weighted imaging (DWI) acquisition.

MRI data were acquired at 3T on a Siemens Skyra with a 20-channel head coil receiver (Erlangen, Germany). Echo planar imaging DWI data were acquired with 60 axial slices, TR/TE = 8400/110 ms, 2.5 mm isotropic voxels, 64 diffusion-weighted images (b=3000 s/mm^2^) and at least 1 b=0 image. A reverse phase-encoded b=0 image was acquired in all cases to correct for B0-field inhomogeneities.

Isotropic T1-weighted magnetization-prepared acquisition gradient echo (MPRAGE) images were also acquired from all participants with the following parameters: TR/TE = 1900/2.5 ms, inversion time = 900 ms, flip angle = 9°, voxel size = 0.9 mm^3^, acquisition matrix 256 × 256 × 192. Intracranial volume was computed from T1 images using SPM12^45^.

### Diffusion-weighted image processing

All DWI data were pre-processed and analysed using MRtrix3^46^, using commands implemented in MRtrix3, or using MRtrix3 scripts that interfaced with external software packages.

Pre-processing of diffusion-weighted images included denoising^47^ and Gibbs ringing removal^48^. Following this, eddy-current and motion correction was performed using FSL’s “eddy” tool (version 6.0). Susceptibility-induced off-resonance fields were estimated using the different phase-encoded b=0 images^49^, after which gross subject movement and eddy-current induced distortions were estimated^50^. The quality of the diffusion datasets was assessed using eddy QC tools^51^. Estimates of subject motion obtained from eddy QC were used to determine whether groups differed in motion. Bias field correction was then performed^52^, and diffusion-weighted images were finally upsampled to a voxel size of 1.3 mm^2^ using cubic b-spline interpolation^53^.

Following these pre-processing steps, fibre orientation distribution (FOD) functions were computed using Single-Shell, 3-Tissue Constrained Spherical Deconvolution (SS3T-CSD^54^), with group-averaged response functions for white matter, grey matter and CSF, using MRtrix3Tissue (http://3Tissue.github.io). Joint bias field and intensity normalisation were then performed.

A study-specific population template image was generated using FOD images from a subset of participants who were randomly selected from the study cohort. FOD images from 40 participants (10 acute mTBI, 10 subacute mTBI, 20 controls) were used to generate a population template image using an iterative registration and averaging approach^55^. FOD images from all subjects were then registered to the unbiased template image using FOD-guided non-linear registration^55,56^.

A whole-brain tractogram was generated using probabilistic tractography on the population template. Twenty million streamlines were generated, which were subsequently filtered to 2 million streamlines to reduce reconstruction biases using the SIFT algorithm^57^.

### Fixel-based analysis

Abnormalities in white matter pathways were assessed using the fixel-based analysis (FBA) framework^36^, whereby the term ‘fixel’ refers to a specific *<u>fi</u>*bre population within a vo*<u>xel</u>*. In brief, this involved computing measures of apparent fibre density (FD), fibre bundle cross-section (FC), and a combined metric of fibre density and cross-section (FDC) for each subject at each white matter fixel in template space.

FD was computed according to the Apparent Fibre Density framework^53^, whereby a quantitative measure of fibre density can be derived from FOD images, as the integral of the FOD along a given direction is proportional to the intra-axonal volume of axons aligned in that direction. FD values for each subject at each fixel were assigned to the template fixel mask so that FD could be compared across groups in corresponding fixels. FC was computed for each fixel by using the non-linear warps to compute the change in FC required to normalise each subject to the template image. While the FD measure provides insight into the intra-axonal density of fibre pathways in a given direction (a microstructural change), the FC measure estimates changes to the spatial extent occupied by a fibre bundle (i.e., a macrostructural or morphological change)^27,36^. Finally, the FDC metric combined both sources of information (FD and FC), to estimate both density and cross-sectional changes in specific white matter pathways.

### Cognitive assessment

Cognitive testing was performed at the time of the MRI scan using CogSport, a computerised cognitive test battery (CogState Ltd, Melbourne, Victoria, Australia). The battery comprised of four tasks: simple reaction time, complex reaction time, one-back, and continuous learning, which have previously been shown to be sensitive to the cognitive effects of concussion in sport^58,59^. These tasks are designed to assess psychomotor function, decision making, working memory, and learning, respectively. Normalised scores from these tests based on neurologically normal healthy individuals^60^ (where 100 reflects the mean score from healthy individuals) were used in subsequent regression analyses to examine the relationship between fibre-specific changes and cognition.

### Statistical analysis

#### Whole-brain fixel-based analysis

Whole-brain fixel-based analysis was performed to compare measures of FD, FC, and FDC in the three groups of interest (acute mTBI, subacute mTBI and controls). Statistical comparisons of these fixel-based metrics were performed between groups at each white matter fixel using a General Linear Model, by performing the following group-wise comparisons: (i) acute mTBI versus controls; (ii) subacute mTBI versus controls; and (iii) acute mTBI versus subacute mTBI. Age and intracranial volume were included as nuisance covariates, but not scanner type due to the unbalanced number of participants on the two scanner types. Connectivity-based smoothing and statistical inference were performed using connectivity-based fixel enhancement (CFE), using 2 million streamlines from the template tractogram and default smoothing parameters (smoothing = 10 mm full-width half-maximum)^61^. Family-wise error (FWE)-corrected p-values were assigned to each fixel using non-parametric permutation testing over 5000 permutations^62^. Statistical significance was set at an FWE-corrected p-value < 0.05.

#### Tract-of-interest regression analyses

The fixels that exhibited significant differences in our whole-brain fixel based analyses were selected for exploratory post-hoc tract-of-interest analyses, to examine if the observed fixel-based differences were associated with clinical measures.

#### Association with clinical measures

For post-hoc analyses with clinical measures, there were five fixel clusters selected: the genu, splenium, and body of the corpus callosum, left frontal white matter (WM) cluster, and left parahippocampal/isthmus WM cluster. These fixel clusters were selected from significant results from whole-brain FBA: between the acute mTBI and control groups for the genu and left parahippocampal/isthmus cluster; and between the acute and subacute mTBI groups for the remaining three fixel clusters (the splenium, the body of corpus callosum, and left frontal WM cluster). Mean FD, FC and/or FDC values were computed within each of these five tract clusters, for the fixel-based metrics that exhibited a significant difference upon whole-brain FBA (i.e., if significant differences were only observed in a given cluster for the FD metric, only FD was examined for these post-hoc analyses). Given that post-hoc regression analyses were performed only in the mTBI participants, we then expressed FD, FC, and FDC values as a percentage change in the mTBI participants from the control group mean.

Exploratory post-hoc linear models were performed within the acute mTBI cohort, to examine the relationship between mean FD/FC/FDC in the five fixel clusters with time since injury. Intracranial volume was included as a covariate in these analyses (given the likely association between FC and head size, and inherent interdependency of FD, FC, and FDC), and Bonferroni correction used to correct for the five tract comparisons.

We then explored the relationship between mean FD/FC/FDC in the five fixel clusters and cognitive scores across the mTBI cohort (both acute and subacute mTBI). Here, linear models were performed to examine the relationship between mean fixel-based metrics within each of the five fixel clusters, and normative scores (expressed as a percentage of the mean value from a healthy control sample) for the four cognitive tests (assessing psychomotor function, decision making, working memory, and learning). Intracranial volume was again included as a covariate in these analyses, and Bonferroni correction used to correct for the five tract comparisons.

All post-hoc statistical analyses were performed in R (version 3.6.3).

### Data availability

The data that support the findings of this study are available upon reasonable request from the corresponding author. The data are not publicly available as they include participant data that could compromise the privacy of participants.

## Results

### Participants

Participant demographics are available in Table 1. The professional athletes in the acute mTBI group had a mean age (± SD) of 24.07 (± 3.82), and had all been scanned within 12 days of injury, with a mean delay since the injury of 7.6 days (± 3.3) [range: 3-12 days]. Athletes in the subacute mTBI group had a mean age of 25.18 (± 3.45), and had all been scanned later than two weeks but within 3 months of injury, with a mean delay between MRI scan and injury of 34.33 days (± 19.4) [range: 16-86 days]. There were no significant differences between the three groups in age, intracranial volume, or DWI motion, although the subacute mTBI group had marginally higher intracranial volume than the acute mTBI and control groups (Table 1).

Athletes in the acute and subacute mTBI groups had comparable years of professional sporting career, and did not differ substantially in CogSport cognitive test measures at the time of MRI scan (Table 1).

### Whole-brain fixel-based analysis

Figure 1 shows the fixel-wise pattern of fibre density and cross-section (FDC) differences when comparing the three groups (acute mTBI, subacute mTBI, and control groups), coloured by family-wise error (FWE)-corrected p-value. While increased FDC was observed in the acute mTBI group when compared to both the control and subacute mTBI groups, predominantly in callosal fibre structures, there was no evidence of decreased FDC in the acute mTBI group when compared to the other groups. In contrast, we did not observe any significant increases in FDC in the subacute mTBI group when compared to controls or acute mTBI. Some fibre structures appeared to have lowered FDC in the subacute mTBI group (namely, the left fornix); however, these did not survive FWE correction.

**Figure 1:**
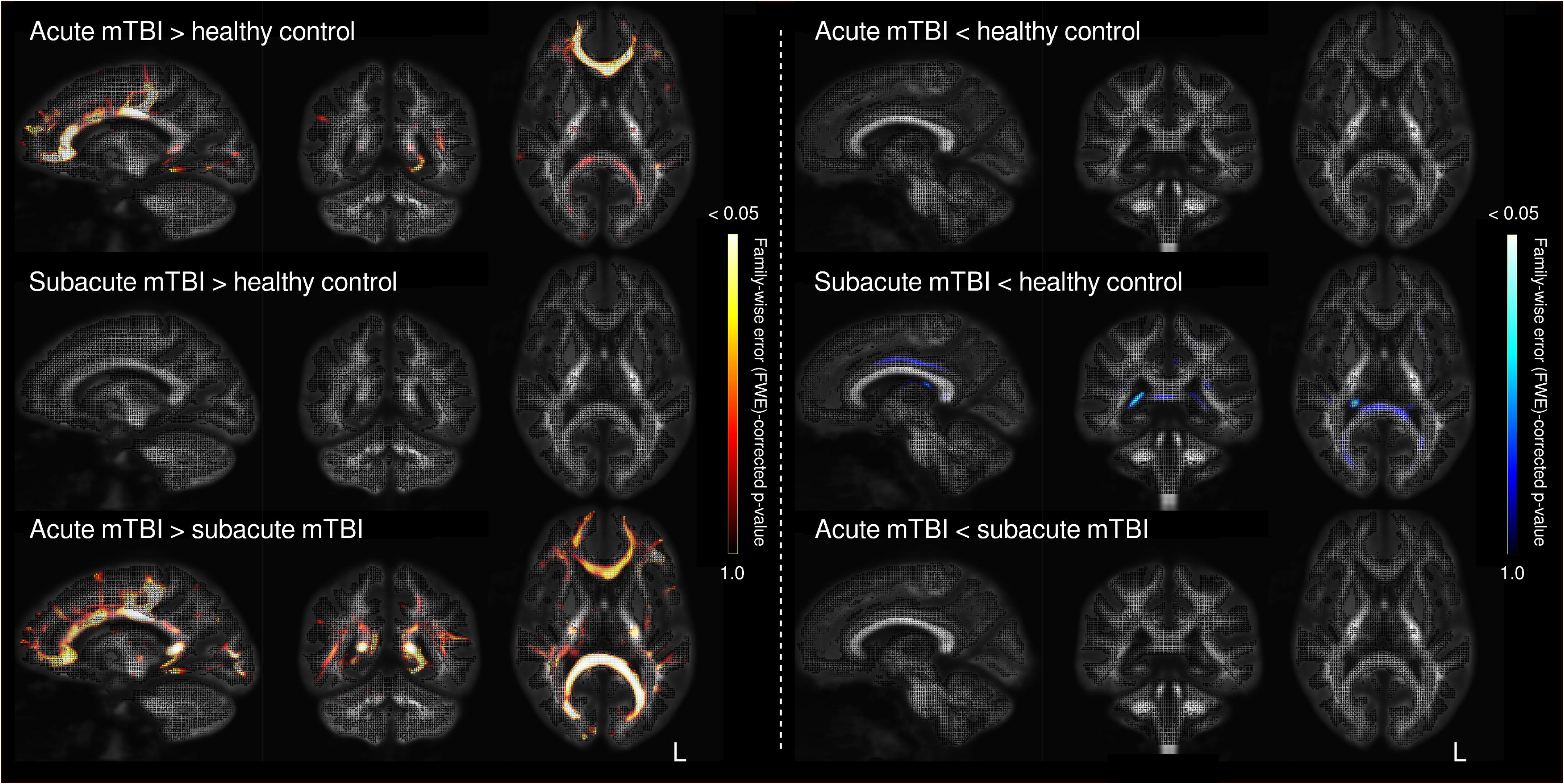
Fixel-based analysis results for fibre density and cross-section in acute and subacute mTBI. Whole-brain fixel-based analysis was performed to compare the acute mTBI, subacute mTBI and neurologically healthy control groups. The results of these whole-brain analyses are shown for the fibre density and cross-section (FDC) metric, on a single sagittal, coronal and axial slice. Fixels (specific fibre populations within a voxel) are unthresholded and coloured by family-wise error (FWE) corrected p-value. As can be appreciated from these comparisons, increased FDC was observed in the acute mTBI cohort, when compared to both the control and subacute mTBI groups, particularly in callosal fibre structures. In contrast, subtle decreases in FDC were apparent in the subacute, but not acute, mTBI group, although these did not survive multiple comparison correction.

Participants with acute mTBI had a small number of fixels with a statistically significant increase in fibre density (FD) compared to controls, in the left posterior parahippocampal white matter extending into the isthmus (FWE-corrected p-value < 0.05). A significant increase in FDC was also observed in the acute mTBI group compared to controls in the genu and body of the corpus callosum (Figure 2).

**Figure 2:**
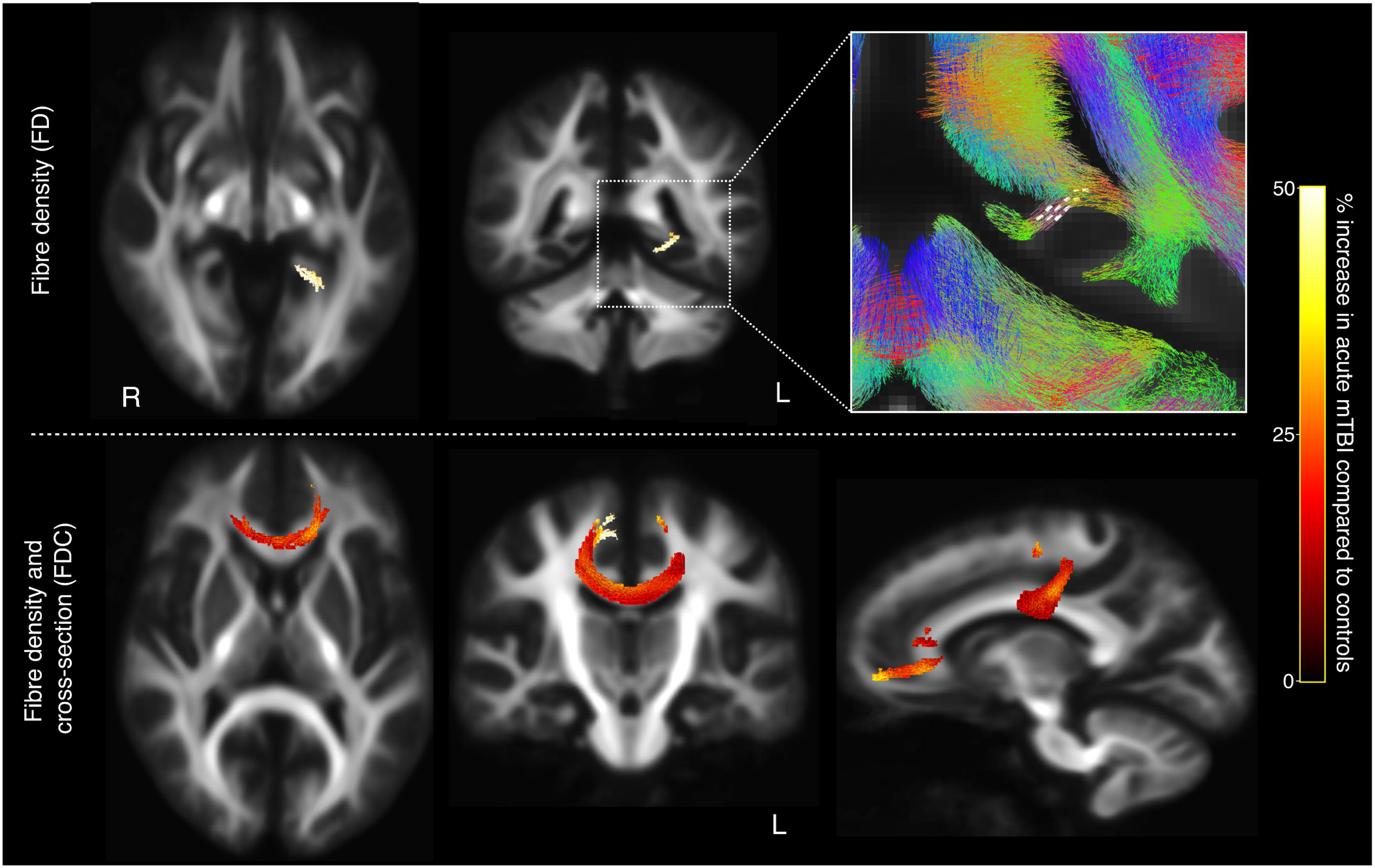
Fibre tracts exhibiting significant abnormalities in acute mTBI. Top row: acute mTBI participants exhibited significant (FWE-corrected p-value < 0.05) increases in fibre density (FD) within select fixels in the left parahippocampal white matter. These fixels are mapped onto the template tractogram and shown on axial and coronal slices at 10 mm increments and are coloured by their percentage increase in the acute mTBI group compared to controls. The inset on the right shows the affected fixels (white lines), with the template tractogram overlaid. Bottom row: significant increases in the fibre density and cross-section (FDC) metric were observed in the acute mTBI group compared to controls, in the genu and body of the corpus callosum. These fixels are again mapped onto the template tractogram, with the axial slice displaying the tract streamlines at 10 mm increments corresponding to the affected fixels in the genu, and the coronal slice displaying the streamlines corresponding to the affected fixels in the body of the corpus callosum. The right image shows a sagittal slice with all affected streamlines displayed.

No significant changes were observed in the subacute mTBI group when compared to controls, in any of the fixel-based metrics.

Fibre-specific increases appeared to be characteristic of the early stages of mTBI, given that significant differences in the fixel-based metrics were observed when comparing those with a recent concussion (≤ 12 days post-concussion; acute mTBI) to those with a longer delay since concussion (> 12 days post-concussion; subacute mTBI). Figure 3 shows the fibre-specific differences between the acute and subacute mTBI groups, for each of the three fixel-based metrics. Significantly greater FD was observed in the acute mTBI group compared to the subacute mTBI group in the splenium of the corpus callosum, as well as in select fixels within the left posterior parahippocampal white matter. Greater FC was also evident in the acute mTBI group in the splenium, as well as in an area of left frontal white matter within the superior longitudinal fasciculus. Significant increases in FDC were observed in the acute mTBI group in the fibre tracts affected by the FD and FC metrics—the splenium and left frontal white matter—as well as in the body of the corpus callosum at the level of the motor cortices.

**Figure 3:**
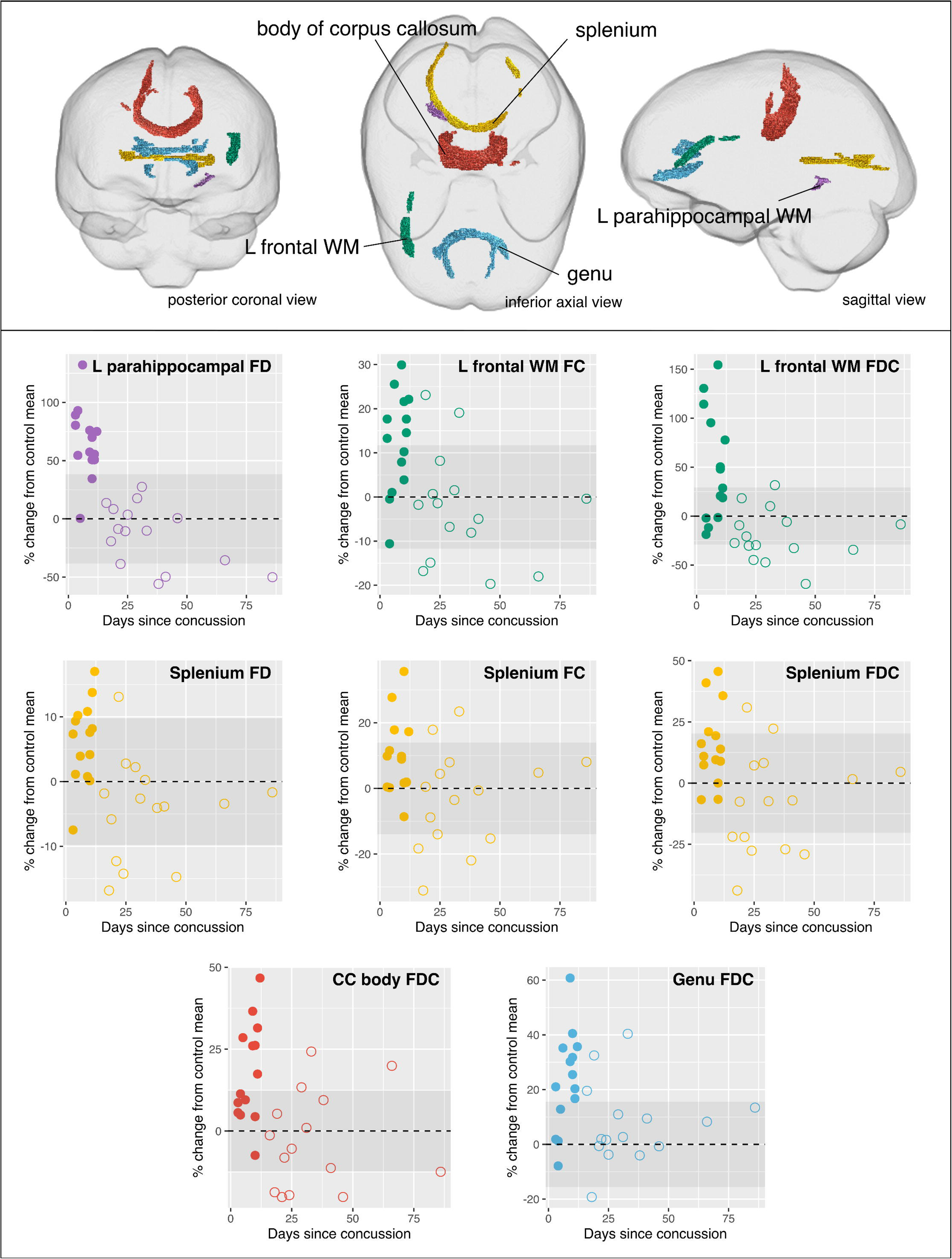
Fibre tracts exhibiting significant differences between the acute and subacute mTBI groups. Fixels exhibiting significant increases in the acute mTBI group when compared to the subacute mTBI group are mapped onto the template tractogram. Significantly affected streamlines are displayed for fibre density (FD; top), fibre cross-section (FC; middle), and fibre density and cross-section (FDC; bottom), and are coloured by the percentage increase in the acute mTBI group compared to the subacute group. For the FD metric, the splenium of the corpus callosum, and the left parahippocampal white matter exhibited significant increases in the acute mTBI group, while for the FC metric, the splenium and the left frontal white matter in the superior longitudinal fasciculus exhibited significant increases. The splenium and body of the corpus callosum, along with the left frontal white matter was affected in the acute mTBI group for FDC metric.

### Tract-of-interest analyses

Fixels that exhibited significant changes in the acute mTBI group when compared to either the control or subacute mTBI groups were grouped into five tract clusters. These tracts-of-interest are shown in Figure 4. Within the acute mTBI group, we did not observe any significant associations between fixel-based metrics in the tracts-of-interest with days since injury (Supplementary Table 1).

**Figure 4:**
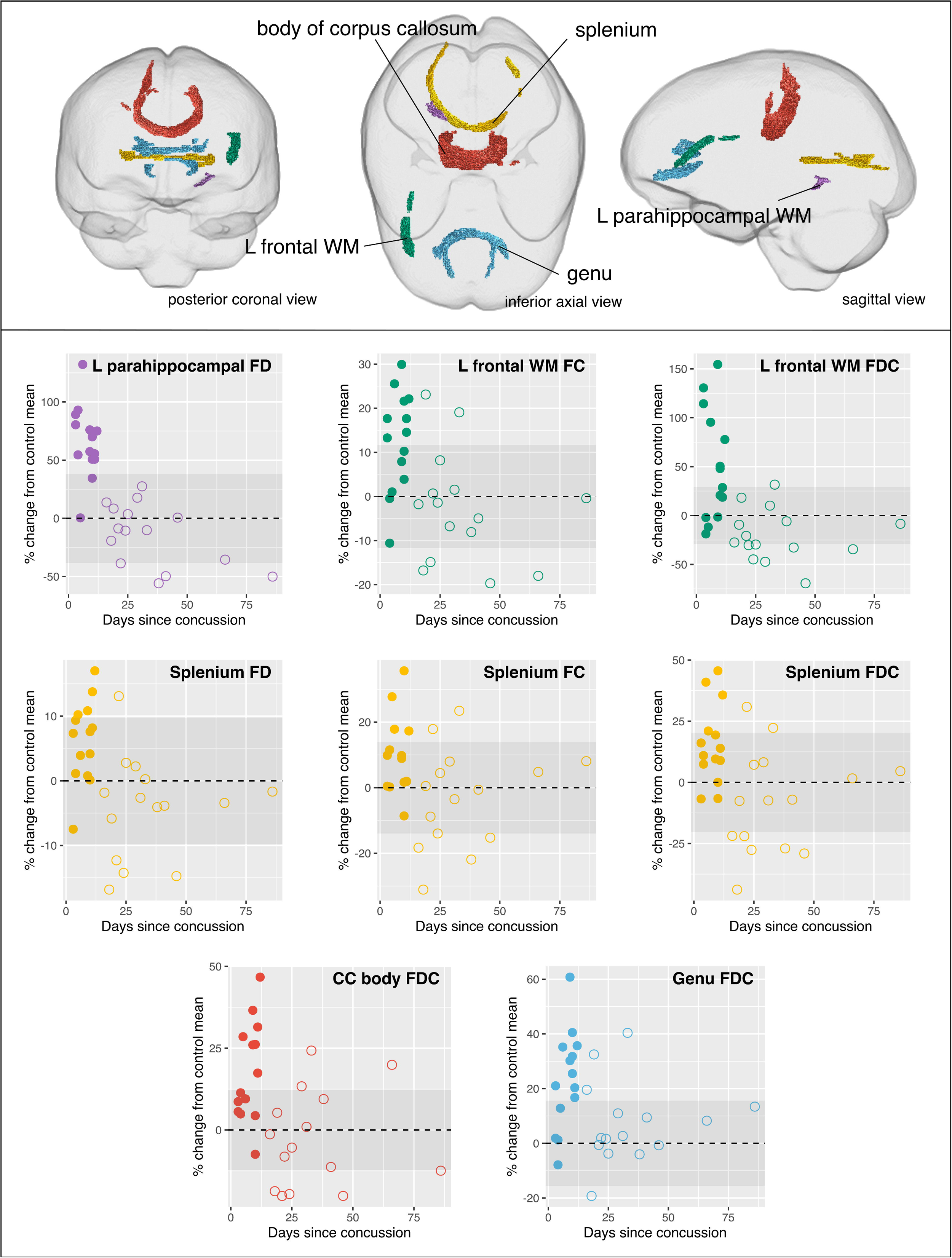
Tracts-of-interest. Top panel: fixels that exhibited significant increases in acute mTBI compared to either controls or subacute mTBI were categorised into five tracts-of-interest. These tracts are shown in coronal, axial, and sagittal glass brain representations, and coloured by the tract to which they belong. Mean fixel-based metrics were computed in these tracts for each mTBI participant for exploratory post-hoc analyses. Bottom panel: scatterplots show the mean fibre density and/or cross-section in each tract of interest, plotted against days since concussion. Data points correspond to individual subjects, with solid circles representing acute mTBI participants, and unfilled circles representing subacute mTBI participants. No significant associations (FDR-corrected *p*-value < 0.05) were observed between fixel-based metrics and days since injury within the acute mTBI group, from linear models including ICV as a covariate in analysis (see Supplementary Table 1). For all scatterplots, fixel-based metrics are expressed as percentage (%) change from the control group mean (the dotted line indicating control mean, and the shaded area showing control group standard deviation).

Cognitive data was available for 27 mTBI participants, while it was unavailable for 2 participants, who were excluded from these analyses. We did not observe any significant associations between mean fixel-based measures in the tracts of interest with cognitive test scores (see Supplementary Table 2). Scatterplots showing the relationship between fixel-based measures in the select tracts and normative cognitive test scores are available in the Supplementary Material (Figures S1-S4).

## Discussion

In this study, we applied advanced diffusion MRI methodology to investigate fibre tract-specific white matter abnormalities in professional athletes after an mTBI. The major findings of our work were that: (i) participants with acute mTBI exhibit abnormalities in selected fibre tracts, which can be interpreted as white matter tract-specific cytotoxic oedema; and (ii) these fibre-specific abnormalities do not seem to persist into the subacute stage beyond 12 days after injury, even when symptoms may persist.

### Acute mTBI is characterised by abnormalities in specific white matter tracts

Abnormalities were observed in specific fibre pathways in acute mTBI participants when compared to control participants in this study, as measured by increases in fibre density and cross-section. Significant changes occurred in the left posterior parahippocampal white matter, and the genu and body of the corpus callosum. Moreover, when comparing acute mTBI to subacute mTBI participants, fibre density and cross-section were also found to be elevated in additional white matter structures: within the splenium of the corpus callosum and left frontal white matter of the superior longitudinal fasciculus.

The location of these tract-specific white matter abnormalities is in line with the body of mTBI literature using DTI, in which certain fibre structures like the corpus callosum have consistently been implicated^11,16,30,63,64^, despite variability in other neuroanatomical locations involved^18,20^. Indeed, callosal structures and midline white matter tracts appear to be preferentially vulnerable to traumatic axonal injury, as has been demonstrated in post-mortem studies^6^, animal models^65^, as well as laboratory reconstructions of sports-related concussions^66^.

In addition to midline structures, we observed significant abnormalities within the left prefrontal white matter and left parahippocampal white matter, but not in any right hemisphere white matter structures. This is consistent with several studies in mTBI^29,64,67^ and TBI more broadly^68,69^ that suggest there is greater left hemisphere abnormality than right. The same left hemisphere fibre tracts that have been shown to exhibit abnormalities in DTI studies in mTBI have also shown leftward asymmetry in healthy individuals^70,71^, whereby higher white matter integrity has been reported. Some have hypothesised that this asymmetry may render these white matter regions more vulnerable to sports-related mTBI^67^. Alternatively, it may be that left-sided injury with verbal disturbance is more often diagnosed as concussion, and overrepresented in these cohorts. Interestingly, in a recent case study from our cohort, increased water content (MRI-T2 relaxometry) was observed within the left frontal white matter, which we suggested reflects selective vulnerability to post-concussive neuroinflammation or oedema in this brain region^72^.

### Increases rather than decreases in fibre density and cross-section in acute mTBI

While there is increasing consensus about which white matter fibre tracts that are affected in mTBI, both increases and decreases in the same DTI-based metrics have been reported, making biological interpretation difficult. In the present study, we observed significant *increases* rather than *decreases* in fibre density and cross-section (a fixel-based, rather than DTI-based metric) in the acute mTBI group. Decreases in this fibre density metric can be interpreted as a loss of axons^36,53^. In contrast, increases in fibre density can be interpreted as an increase in the total intra-axonal volume occupied by fibre structures aligned in a given direction, even in the presence of crossing-fibre pathways^53^. In the present context, our finding of increased FD is entirely compatible with cytotoxic oedema in these fibre structures. An increase in membrane permeability could result in axonal swelling^73–75^, and larger axonal diameters^76^ (see Figure 5). In the presence of enlarged axons, fibre bundles could also increase in cross-section, which would drive the observed increases in the FC metric. While *decreases* in FD have been suggested to reflect acute injury and *decreases* in FC have been suggested to reflect chronic white matter injury^36,37^, in this context, the FD and FC *increases* are likely to reflect the same underlying acute process.

**Figure 5:**
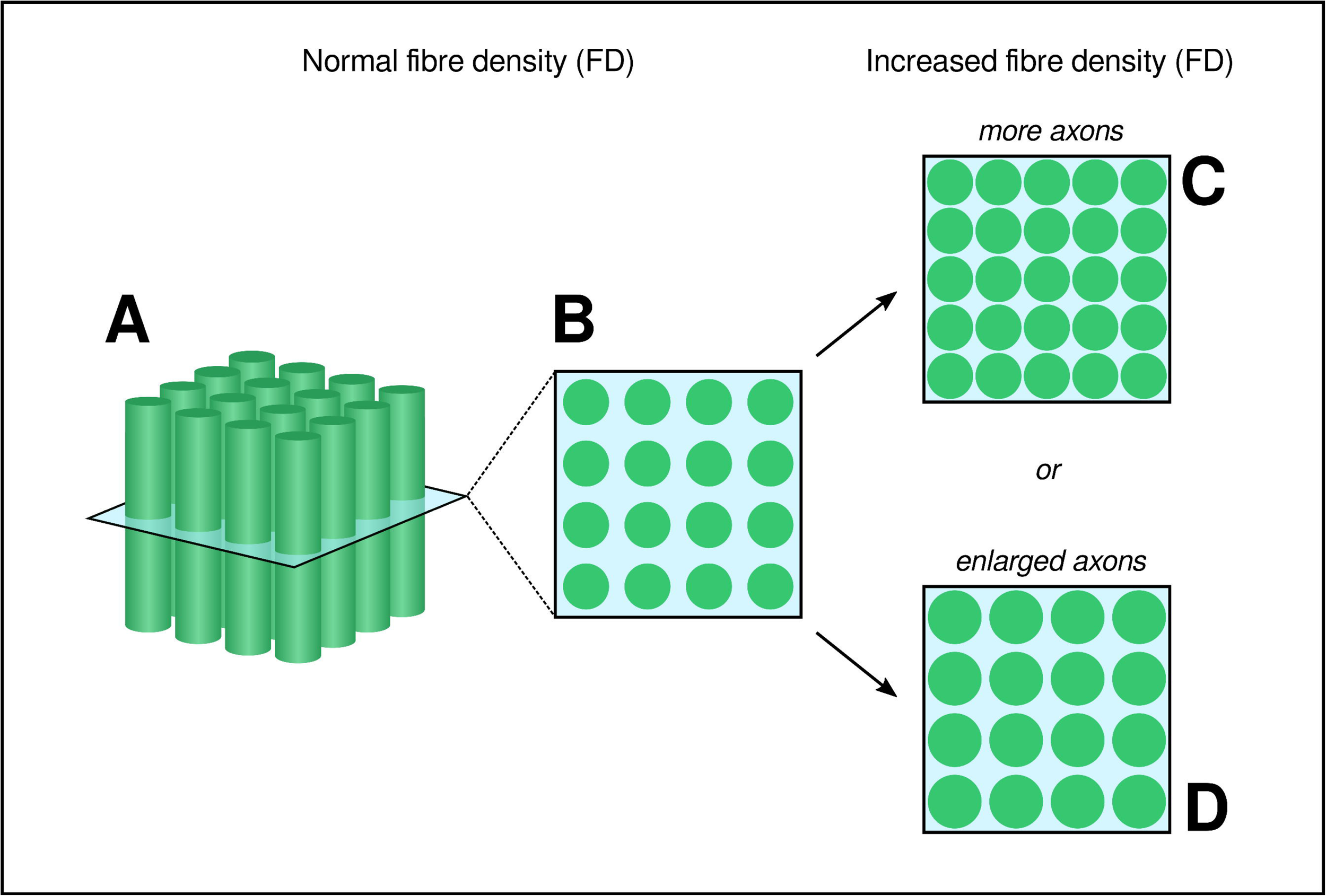
Schematic figure showing potential causes of increased fibre density (FD). (A) and (B) are adapted from Raffelt *et al*. (2012)^53^, where (A) shows a depiction of axons that are coherently ordered within a single imaging voxel, and (B) shows the perpendicular plane through (A). For DWI data with high *b*-values such as those acquired in this study (*b*=3000 s/mm^2^), the fibre density (FD) metric is approximately proportional to the volume of the intracellular component of axons oriented in a given direction (shown in green in B). Increases in the fibre density (FD) value could arise if there is an increase in the volume of the intracellular component; for example, if there are more axons (C), or if there is increased intracellular volume occupied by the same number of axons (D). In the present study, the observed increase in FD is likely to reflect increased axonal volume in the absence of change to the number of axons (i.e. (D)). Provided that axonal diameters stay within the normal range (≤ 6μm), this increase in FD is entirely compatible with cytotoxic oedema, whereby increased permeability of the axonal membrane can result in axonal swelling.

DTI studies in which increased fractional anisotropy has been reported have similarly suggested that mTBI is characterised by cytotoxic oedema^28,77,78^. In longitudinal DTI studies, persistent FA increases (and/or MD decreases) have been observed at the acute, subacute and chronic stages following injury, for up to 6 months^11,14,79^. The chronic nature of increases in FA has raised questions about its interpretability as cytotoxic oedema^79^, particularly if the inflammatory process were to be followed by necrotic cell death^80^. Chronically increased FA, however, may be misleading, as the same direction of FA changes in the acute and chronic stages following mTBI could arise from different processes, for example, the loss of crossing fibre structures in the chronic phase of the injury^11^.

In our study, the increases in fixel-based metrics observed in the acute mTBI group did not appear to persist into the subacute stage. Indeed, if anything, our findings suggest there may be subtle *decreases* in fibre density and cross-section by the subacute stage (Figure 1), given the greater extent of significant fixel-based differences between the acute and subacute mTBI groups than between the acute mTBI and control groups. Such a finding is in line with a meta-analysis of DTI studies in mTBI that suggest the direction of anisotropy changes may relate to timing since injury^22^. This would be compatible with an oedematous process, whereby cytotoxic oedema in the acute stage is followed by axonal loss at later time points. Alternatively, oedema may be followed by recovery of neurons^81^, and indeed faster recovery profiles have been suggested to be characteristic of professional athletes who are exposed to repeated concussive events^82^. It is also possible that a given individual could follow one of these two trajectories: (1) recovery following transient inflammation and/or oedema; or (2) cytotoxic oedema followed by neuronal loss due to axotomy. Thus, the trajectory of changes in fixel-based metrics may predict clinical recovery in that individual.

### The clinical significance of fibre-specific increases in acute mTBI

There is some evidence from DTI studies to suggest that inflammatory processes in acute mTBI may relate to symptom severity and clinical outcomes^28,77,78^. In the present study, while we hypothesised that fibre density and cross-sectional increases observed in the acute mTBI group would relate to cognitive abnormalities, we did not find any evidence to support this.

Cognitive abnormalities and symptom severity following mTBI are generally quite transient, with longitudinal studies showing abnormal clinical measures evident at 24 to 48 hours normalising by 8 or 9 days following concussion^13,14,16^. Given that our acute mTBI group had a mean time since the injury of 7.5 days (Table 1), it is likely that any cognitive abnormalities that may have been evident in this group would have normalised by the time of our clinical assessment. This may explain why we did not observe any significant associations between the fibre-specific abnormalities in the acute mTBI group with reduced cognitive scores. It should be noted that in DTI studies, clinical signs and symptoms, rather than cognitive performance, have been shown to be more sensitively related to brain injury, as measured by increases in fractional anisotropy^28,78^.

Within the acute mTBI group, we also did not find any evidence of an association between tract-specific abnormalities and days since injury. It is possible that we were under-powered to observe such an association within the acute mTBI group. Moreover, it is difficult to assess the relationship of these tract-specific changes with time in a cross-sectional study design. Nonetheless, given that tract-specific measures were not markedly elevated in individuals at the subacute stage of injury, these tract-specific increases could be considered a useful marker of the early stage (first 12 days) of mTBI, irrespective of time since injury.

In our study, the acute period was relatively long (≤ 12 days). This period was selected to align with current protocols for the AFL footballers (a minimum 12-day break from return-to-play following a concussion^44^). Our present findings suggest that tract-specific abnormalities may normalise after 12 days. However, it is essential to emphasise the highly individualised nature of brain injury, symptoms and recovery. Injured players may exhibit different recovery profiles and require an individualised assessment to guide return-to-play.

### Technical advantages of fixel-based analysis

One of the major advantages of applying fixel-based analysis to investigate mTBI is that its fibre-specific nature offers greater interpretability than more commonly used approaches such as DTI. Changes in DTI-based metrics such as fractional anisotropy and mean diffusivity are potentially attributable to a range of non-specific causes, including partial volume effects^83^, loss of crossing-fibre populations^25,27^, and gliosis^84^. This can result in difficulty interpreting the clinical significance of DTI-based findings. In the present study, due to how diffusion MRI data have been acquired, modelled, and analysed, we can better interpret changes in fibre density and cross-section (see Figure 5).

While DTI has been applied to detect axonal injury in individuals with moderate to severe TBI^85^, no neuroimaging tools currently exist that can diagnose mTBI in individuals^21^. It has been suggested that DTI possesses adequate sensitivity to detect white matter abnormalities in mTBI at both acute and subacute stages using group analyses^18^; however, the inherent biases introduced by voxel-based analyses and tract-based spatial statistics (TBSS) make their use for detection of mTBI in individual subjects inappropriate^86^. In this regard, fixel-based analysis may prove valuable, as it enables the detection of highly localised, yet important tract-specific changes–such as those observed here in the left parahippocampal white matter–without being confounded by the presence of crossing-fibre populations. Moreover, if fibre-specific changes follow a dynamic trajectory over time (as our cross-sectional findings suggest), FBA may be valuable in guiding safe return-to-play following injury in the future.

It should be noted that there have been various advanced diffusion methods that have been developed over the past decade. Some mTBI studies have implemented such diffusion models including diffusion kurtosis imaging (DKI) and neurite orientation dispersion and density imaging (NODDI)^14,87–91^. However, the major advantage of FBA is that it moves entirely beyond a tensor-based model in which comparisons are inherently voxel-averaged, to using a fibre tract-specific model which enables more directly interpretable measures of structural connectivity^36^. FBA has now been applied across numerous neurological disorders^38^, as well as in recent TBI studies^92–94^. Of note, our findings corroborate recent work in a small cohort of Australian Rules footballers that has similarly reported increases in fixel-based metrics in concussed individuals^95^.

### Limitations and future directions

While we argue that FBA presents a major technical advantage, there are limitations pertaining to our study cohort. Our findings are based on a relatively small cohort of mTBI participants, and although we had a well-matched sample of control participants, demographic measures that could influence fibre-specific measures (such as years of education) were not available for these participants. Our study included a highly controlled cohort of professional athletes, in whom findings may not be generalisable to other TBI cohorts.

Professional AFL players often report numerous concussions during their sporting careers^72^, and there is increasing evidence regarding the deleterious effects of repeat concussions^96,97^. However, in this study, we were unable to account for the number of previous concussions, as this self-reported and often unreliable data was not available for all participants. Despite this, the subacute mTBI cohort used in this study can in many ways be considered an almost ideal control cohort for the acute mTBI participants, given that they were professional athletes likely to be well-matched in demographic measures, concussion history, sporting ability and training requirements. The similarities in the affected fibre pathways when comparing the acute and subacute mTBI groups, to when we compared the acute mTBI to healthy control cohort, provides comfort that these changes relate to acute effects of mTBI, and not to other potential differences between the acute and healthy control participants.

Finally, while we hypothesise on the potential trajectory of fibre-specific changes over the acute and subacute stages of mTBI, we cannot assess the temporal changes with our cross-sectional data. Given that this study included professional athletes who were recruited by team doctors, their inclusion in the study was potentially driven by concern from the medical professionals who assessed their recovery and was also only possible when athletes did not have substantial training and competition commitments. The acute mTBI group represented a cohort of professional athletes recruited during their mandated rest period. However, participants in the subacute mTBI group (more than 12 days after injury) were likely recruited due to poor recovery or persistent symptomatology. The subtle abnormalities detected in the subacute stage in our cohort may be characteristic to those with poor recovery or persistent symptoms (Figure 1). Future work investigating the longitudinal trajectory of fibre-specific changes across acute and subacute stages will be invaluable in this regard.

## Conclusions

Acute mTBI is characterised by subtle abnormality in select fibre structures. The location and direction of these fibre-specific changes are consistent with previous literature, and our findings support the theory of cytotoxic oedema in the days to weeks following mTBI. However, we did not find evidence to suggest that these fibre tract changes in the first two weeks following injury are associated with cognitive deficits nor time since injury. Future work incorporating fixel-based analysis into longitudinal studies will be critical for further understanding these changes including the time course of these fibre tract changes following sports-related mTBI.

## Abbreviations

AFL: Australian Football League
DTI: diffusion tensor imaging
DWI: diffusion-weighted imaging
FBA: fixel-based analysis
FA: fractional anisotropy
FC: fibre bundle cross-section
FD: fibre density
FDC: fibre density and cross-section
FOD: fibre orientation distribution
MD: mean diffusivity
mTBI: mild Traumatic Brain Injury

## Acknowledgements

The authors wish to thank the Australian Football League (AFL) Doctors Association, AFL Players Association, Dr Patrick Clifton (former AFL Operations and Innovations Manager) and Dr Peter Harcourt (former AFL Medical Director) for their support of the project. We also thank all the recruited participants who gave their valuable time to participate in this study. The Florey Institute of Neuroscience and Mental Health acknowledges the strong support from the Operational Infrastructure Support Grant. We also acknowledge the facilities, and the scientific and technical National Collaborative Research Infrastructure Strategy (NCRIS) capability at the Florey node, and the Victorian Biomedical Imaging Capability (VBIC).

## Author contributions

**Remika Mito:** Conceptualization, Formal analysis, Visualization, Writing – Original Draft; **Donna Parker:** Investigation, Project administration, Data curation, Writing – Review & Editing; **David Abbott:** Resources, Project administration, Funding acquisition, Writing – Review & Editing; **Michael Makdissi:** Investigation, Resources, Project administration, Data curation, Funding acquisition, Writing – Review & Editing; **Mangor Pedersen:** Conceptualization, Writing – Review & Editing, Supervision; **Graeme Jackson:** Conceptualization, Funding acquisition, Writing – Review & Editing, Supervision.

## Funding

This study was supported in part by the National Health and Medical Research Council (NHMRC) of Australia (G.J. Practitioner Fellowship; grant number #1060312). R.M. is supported by grants from Brain Australia and the Brain Foundation. D.A. is supported by fellowship funding from the National Imaging Facility. M.P. is supported by an Emerging Grant Fellowship from the Health Research Council. Funding for the MRI scans was provided by the AFL.

## Competing interests

Dr Michael Makdissi is a consultant Sport and Exercise Medicine physician at Olympic Park Sports Medicine Centre and Chief Medical Officer for the Australian Football League (AFL). He is a member of the International Concussion in Sport Group. He has previously received research funding from the AFL and non-financial research support from CogState Pty Ltd. He has attended meetings organised by the International Olympic Committee, the National Football League (USA), the National Rugby League (Australia), and FIFA (Switzerland); however, has not received any payment, research funding, or other monies from these groups other than for travel costs. He is an honorary member of concussion working/advisory groups for the Australian Rugby Union and World Rugby. The remaining authors report no competing interests.

## Supplementary Material

**Supplementary Table 1:** Association between tract FBA metrics and days since injury in acute mTBI participants.

|  | **t-value** | **p-value** |
| --- | --- | --- |
| **Left parahippocampal WM (FD)** | -0.744 | 0.472 |
| **Left prefrontal WM (FC)** | 1.416 | 0.184 |
| **Left prefrontal WM (FDC)** | -0.509 | 0.621 |
| **Body of corpus callosum (FDC)** | 1.910 | 0.082 |
| **Genu of corpus callosum (FDC)** | 2.565 | 0.026 |
| **Splenium of corpus callosum (FD)** | 1.711 | 0.115 |
| **Splenium of corpus callosum (FC)** | -0.228 | 0.824 |
| **Splenium of corpus callosum (FDC)** | 0.443 | 0.666 |

Cells show t-value and p-value from linear models examining the relationship between tract-specific fibre density (FD), fibre cross-section (FC), and fibre density and cross-section (FDC) measures in the five tracts of interest with days since injury in the acute mTBI participants (n = 14). Intracranial volume (ICV) was included as a covariate in all analyses.

**Supplementary Table 2:** Association between tract FBA metrics and cognitive function.

|  | **Psychomotor**<br>**function** | **Decision**<br>**making** | **Working**<br>**Memory** | **Learning** |
| --- | --- | --- | --- | --- |
| **Left**<br>**parahippocampal**<br>**WM (FD)** | t = 0.974,<br>p = 0.340 | t = 1.450,<br>p = 0.160 | t = 0.871,<br>p = 0.392 | t = 0.185,<br>p = 0.855 |
| **Left prefrontal**<br>**WM (FC)** | t = 0.145,<br>p = 0.886 | t = 0.640,<br>p = 0.528 | t = -0.331,<br>p = 0.744 | t = -1.582,<br>p = 0.127 |
| **Left prefrontal**<br>**WM (FDC)** | t = 0.382,<br>p = 0.706 | t = 1.157,<br>p = 0.259 | t = 0.346,<br>p = 0.732 | t = -1.214,<br>p = 0.237 |
| **Body of corpus**<br>**callosum (FDC)** | t = 0.097,<br>p = 0.923 | t = 0.241,<br>p = 0.811 | t = 0.559,<br>p = 0.581 | t = -0.938,<br>p = 0.358 |
| **Genu of corpus**<br>**callosum (FDC)** | t = -0.402,<br>p = 0.692 | t = 0.429,<br>p = 0.671 | t = 0.057,<br>p = 0.955 | t = -1.412,<br>p = 0.171 |
| **Splenium of**<br>**corpus callosum**<br>**(FD)** | t = 1.808,<br>p = 0.084 | t = 2.127,<br>p = 0.0439 | t = 1.436,<br>p = 0.164 | t = 0.147,<br>p = 0.884 |
| **Splenium of**<br>**corpus callosum**<br>**(FC)** | t = 0.764,<br>p = 0.452 | t = 1.361,<br>p = 0.186 | t = 1.295,<br>p = 0.208 | t = 0.215,<br>p = 0.832 |
| **Splenium of**<br>**corpus callosum**<br>**(FDC)** | t = 1.239,<br>p = 0.228 | t = 1.786,<br>p = 0.087 | t = 1.471,<br>p = 0.154 | t = 0.166,<br>p = 0.869 |

Cells show t-value and p-value from linear models examining the relationship between tract-specific fibre density (FD), fibre cross-section (FC), or fibre-density and cross-section (FDC) measures in the five tracts of interest with cognitive measures. Intracranial volume (ICV) was included as a covariate in all analyses.

**Supplementary Figure 1:**
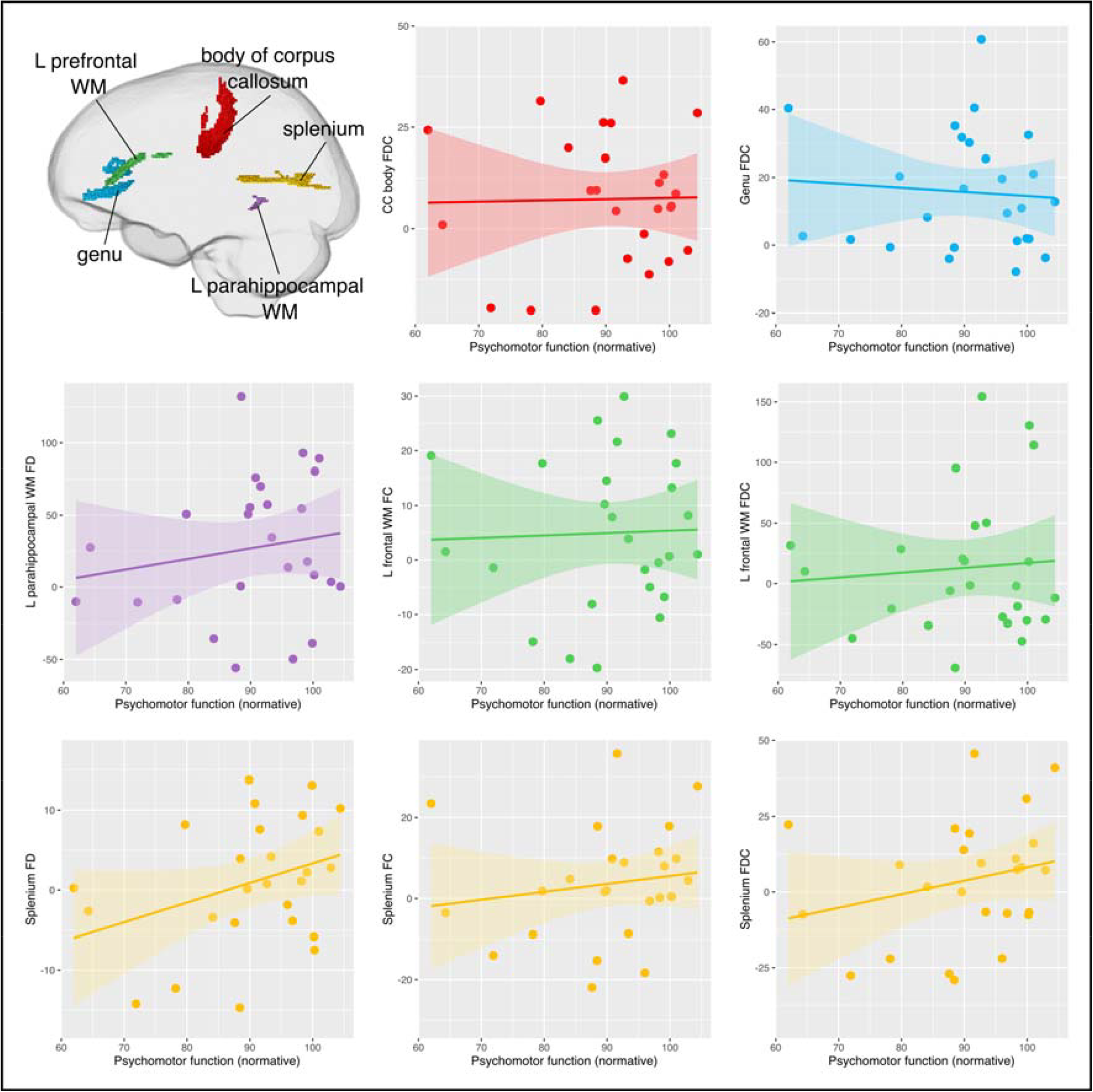
Scatterplots showing the relationship between simple reaction time score (psychomotor function) and fixel-based measures in tracts-of-interest. Mean fixel-based metrics were computed in each tract of interest (glass brain at top left shows included tracts) for each mTBI participant, and expressed as a percentage change from the healthy control mean. No significant relationships were observed between psychomotor function and tract FD. Simple reaction time (psychomotor function) scores are expressed as normative values, where 100 reflects the average value from a healthy sample. Intracranial volume (ICV) was included in linear models for statistical analysis.

**Supplementary Figure 2:**
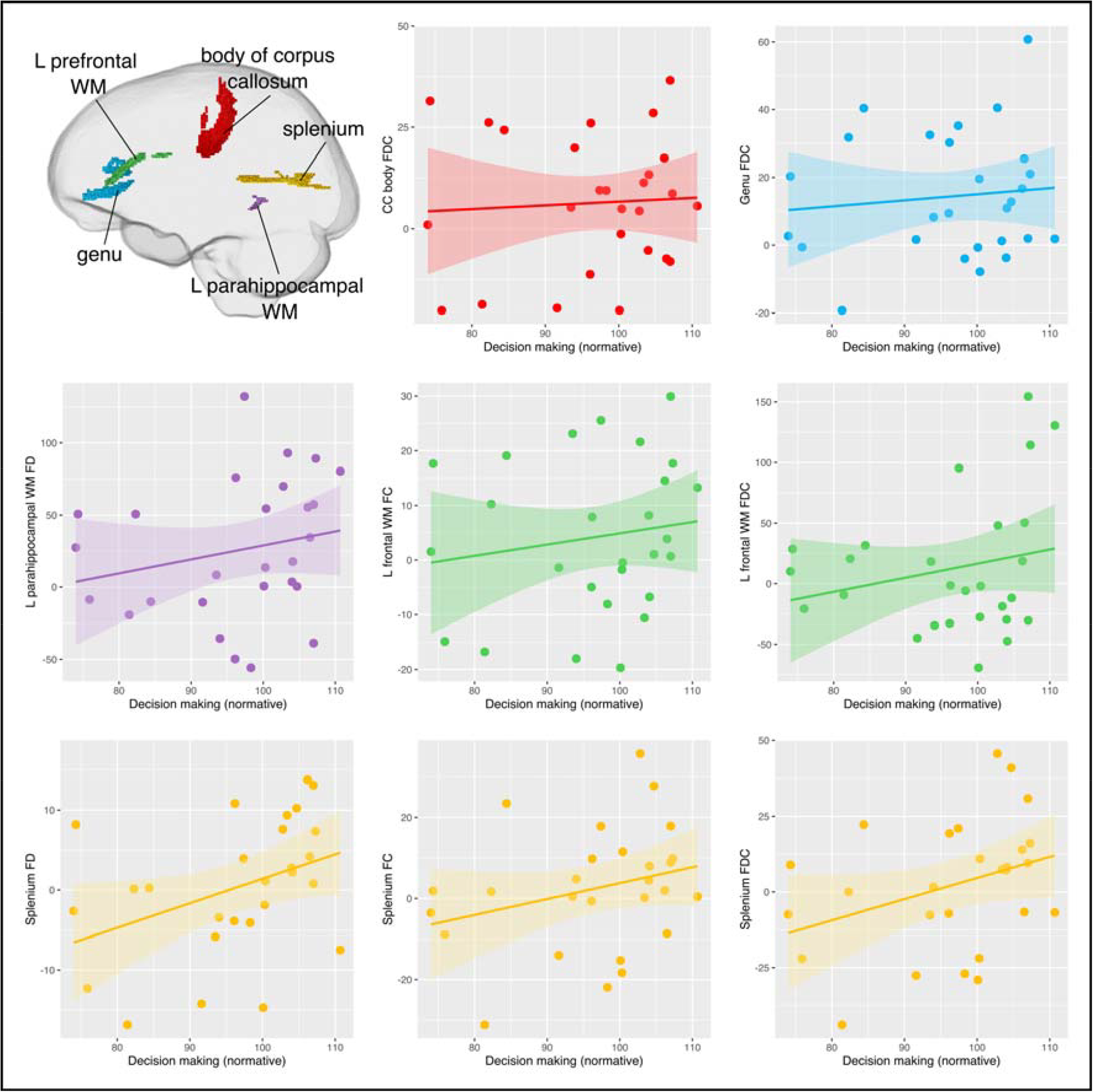
Scatterplots showing the relationship between complex reaction time score (decision making) and fixel-based measures in tracts-of-interest. Mean fixel-based metrics were computed in each tract of interest (glass brain at top left shows included tracts) for each mTBI participant, and expressed as a percentage change from the healthy control mean. No significant relationships were observed between decision making scores and mean tract-based measures, although there was a strong association between decision making and fibre density (FD) in the splenium of the corpus callosum. Decision making scores are again expressed as normative values, where 100 reflects the average value from a healthy sample. Intracranial volume (ICV) was included in linear models for statistical analysis.

**Supplementary Figure 3:**
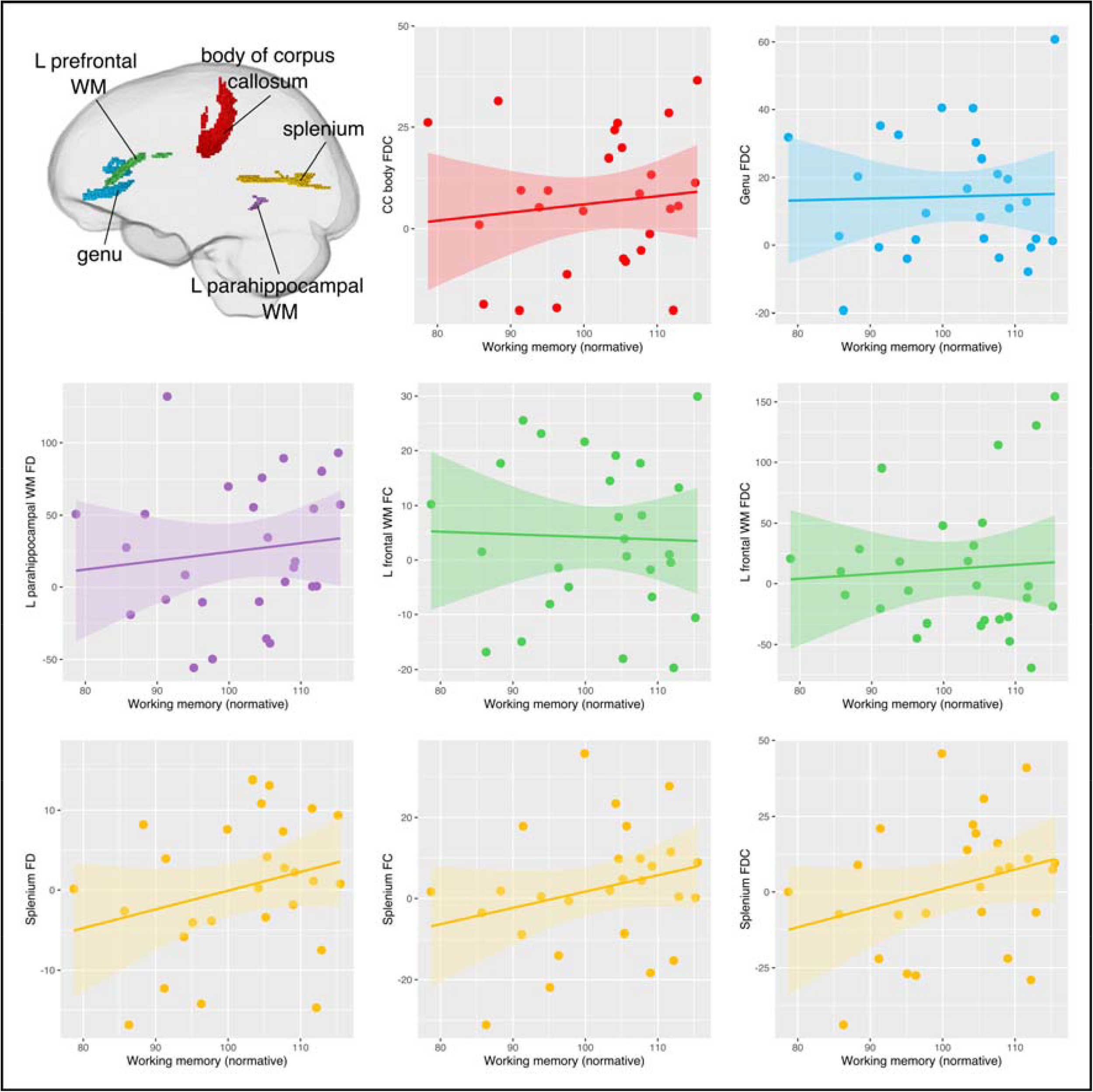
Scatterplots showing the relationship between complex one-back score (working memory) and fixel-based measures in tracts-of-interest. No significant associations were observed between working memory scores and fixel-based measures in any of the tracts-of-interest. One-back (working memory) scores are again expressed in normative values, with 100 corresponding to the average value from a healthy sample.

**Supplementary Figure 4:**
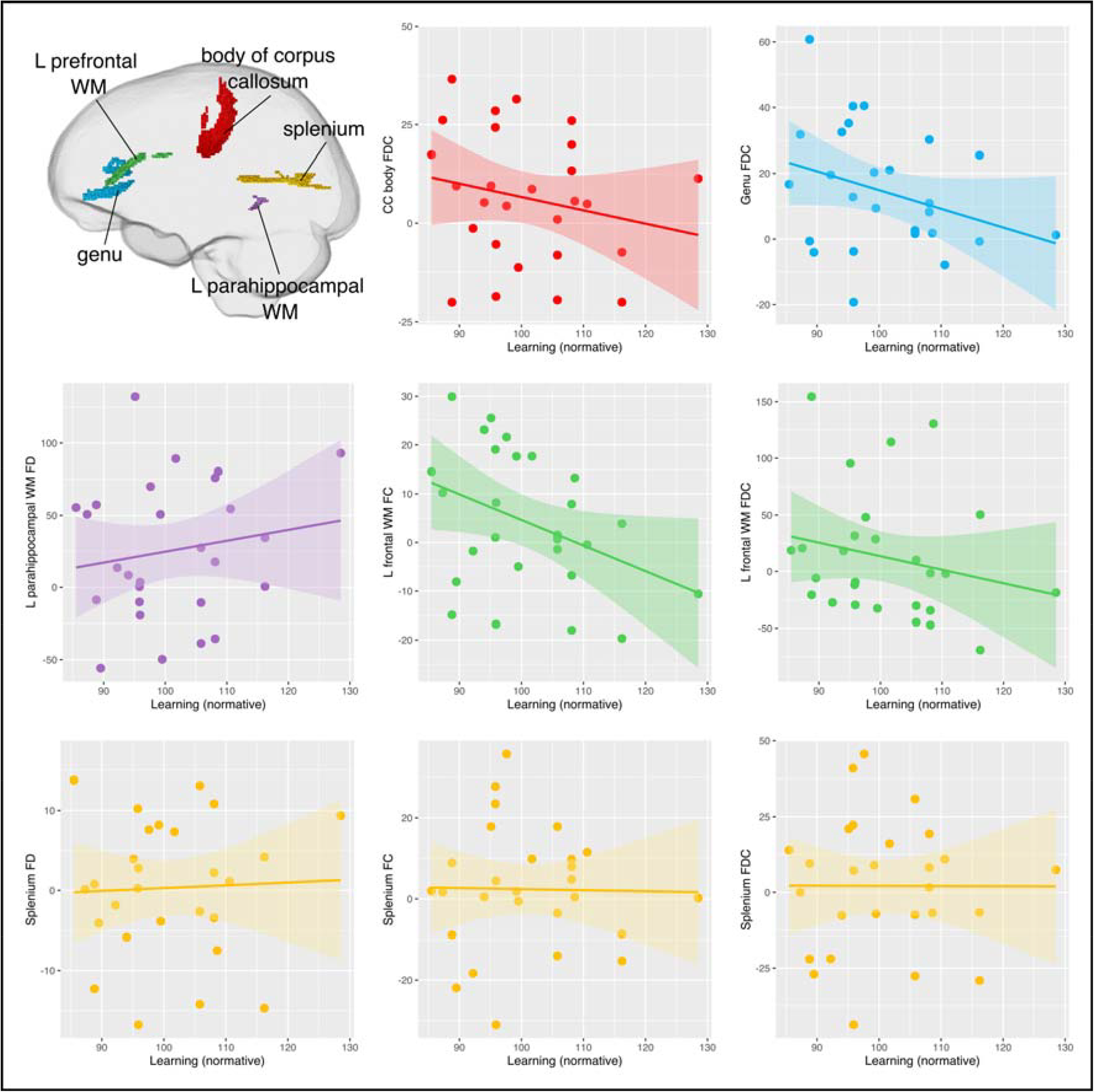
Scatterplots showing the relationship between continuous learning task (learning) and fixel-based measures in tracts-of-interest. No significant relationships were observed between the learning score (again expressed as a normative value) and fixel-based measures in the tracts-of-interest.

## Notes

### Author Declarations

The University of Melbourne Human Research Ethics Committee (ID: 0830367) and Austin Health Human Research Ethics Committee (ID: 49573/2019 and H2012/04475) gave ethical approval for this work.

### Summary of Updates

Title updated. Participants scanned using different scanner and head coil excluded from study, and Figures 1-4 revised with updated results. Supplemental files updated.

